# Risk Factors for ICU Admission, Mechanical Ventilation and Mortality in Hospitalized Patients with COVID-19 in Hubei, China

**DOI:** 10.1101/2020.08.31.20184952

**Authors:** Hong Gang Ren, Xingyi Guo, Kevin Blighe, Fang Zhu, Janet Martin, Luqman Bin Safdar, Pengcheng Yang, Dao Wen Wang, Qinyong Hu, Nan Huo, Justin Stebbing, Davy Cheng

**Affiliations:** Department of Internal Medicine, Tongji Medical College, Huazhong University of Science and Technology, Wuhan, China; Division of Epidemiology, Department of Medicine, Vanderbilt Epidemiology Center, Vanderbilt University School of Medicine, Nashville, USA; Clinical Bioinformatics Research Ltd., London, UK; Department of Anesthesia & Perioperative Medicine, Centre for Medical Evidence, Decision Integrity and Clinical Impact and Department of Epidemiology &Biostatistics, Western University, London, Ontario, Canada; The Key Laboratory of Biology and Genetic Improvement of Oil Crops, Oil Crops Research Institute, Chinese Academy of Agricultural Sciences, The Ministry of Agriculture and Rural Affairs, Wuhan 430062, China; Cancer Center, Renmin Hospital of Wuhan University, No. 99, Zhangzhidong Road, Wuchang District, Wuhan, China; Department of Epidemiology, Health Sciences Research, Mayo Clinic, Rochester, Minnesota, USA; Department of Surgery and Cancer, Division of Cancer, Imperial College London, Hammersmith Hospital Campus, Du Cane Road, London, UK

**Keywords:** COVID-19, CFR, ICU admission, mechanical ventilation, glucocorticoid

## Abstract

**Purpose:** To examine the risk factors for Intensive Care Unit (ICU) admission, mechanical ventilation and mortality in hospitalized patients with COVID-19.

**Methods:** This was a retrospective cohort study including 432 patients with laboratory-confirmed COVID-19 who were admitted to three medical centers in Hubei province from January 1^st^ to April 10^th^ 2020. Primary outcomes included ICU admission, mechanical ventilation and death occurring while hospitalized or within 30 days.

**Results:** Of the 432 confirmed patients, 9.5% were admitted to the ICU, 27.3% required mechanical ventilation, and 33.1% died. Total leukocyte count was higher in survivors compared with those who died (8.9 vs 4.8 × 10^9^/l), but lymphocyte counts were lower (0.6 vs 1.0 × 10^9^/l). D-dimer was significantly higher in patients who died compared to survivors (6.0ug/l vs 1.0ug/l, p<0.0001. This was also seen when comparing mechanically versus non-mechanically-ventilated patients. Other significant differences were seen in AST, ALT, LDH, total bilirubin and creating kinase. The following were associated with increased odds of death: age > 65 years (adjusted hazard ratio (HR 2.09, 95% CI 1.02-4.05), severe disease at baseline (5.02, 2.05-12.29), current smoker (1.67, 1.37-2.02), temperature >39^°^ C at baseline (2.68, 1.88-4.23), more than one comorbidity (2.12, 1.62-3.09), bilateral patchy shadowing on chest CT or X-ray (3.74, 1.78-9.62) and organ failure (6.47, 1.97-26.23). The following interventions were associated with higher CFR: glucocorticoids (1.60, 1.04-2.30), ICU admission (4.92, 1.37-17.64) and mechanical ventilation (2.35, 1.14-4.82).

**Conclusion:** Demographics, including age over 65 years, current smoker, diabetes, hypertension, and cerebrovascular disease, were associated with increased risk of mortality. Mortality was also associated with glucocorticoid use, mechanical ventilation and ICU admission.

**Take-Home Message:** COVID-19 patients with risk factors were more likely to be admitted into ICU and more likely to require mechanical ventilation.

## INTRODUCTION

As of May 5 2020, the COVID-19 pandemic has led to over 3.6 million cases of infection and over 270,000 deaths globally, according to the World Health Organization. In earlier reports, the case fatality rate (CFR) of critically ill patients with COVID-19 has been as high as 61.5%^1-11^. COVID19 exhibits distinctive features from acute respiratory distress syndrome (ARDS)^12^ and, although a number of therapeutic options exist, including lopinavir/ritonavir combination, remdesivir, arbidol, convalescent plasma, traditional Chinese medicine, and stem cell infusion remain experimental, there is a lack of evidence regarding their net benefits and risks. To date, there are no specific drugs or therapeutic interventions which have been proven to reduce mortality, nor is the development of an effective vaccine expected this year. However, guidelines from a panel of experts have recently emerged, covering many aspects relating to critical care management of COVID-19 patients^13^.

Currently, few studies have focused on the risk factors for death in hospitalized patients with COVID-19^10-11^. Frailty index (FI) has been examined as a potentially useful measure of outcome^14^. Also, in a study of 191 hospitalized patients, Zhou et al found that older age, higher sequential organ failure assessment score (SOFA), and D-dimer over 1μg/L were independent predictors of death in patients with COVID-19.^11^ A study of 52 critically ill patients with COVID-19 found that increased age, development of ARDS, and mechanical ventilation were associated with mortality. However, early studies on mortality and associated risk factors have been plagued by incomplete data, lack of longitudinal follow-up and small sample size, since many COVID-19 patients remained hospitalised at time of publication.

The objective of this study was to compare patient demographics, clinical characteristics, and management strategies between COVID-19 fatalities and survivors, and to identify risk factors for Intensive Care Unit (ICU) admission, mechanical ventilation and mortality in hospitalized patients with confirmed COVID-19.

## METHODS

### Study population and data sources

This was a retrospective cohort study using electronic medical record data from January 1st to February 29th, 2020 of 432 inpatients with confirmed COVID-19 from three academic medical centers in Hubei province (Xiehe medical center, Hannan medical center, and Enshi medical center). All of these three medical centers have fully functional ICU capabilities. Patients who were diagnosed with COVID-19 before January 1^st^ or after February 29^th^ were excluded due to lack of standardized treatments. Patients with missing data regarding ICU admission or mechanical ventilator use were excluded from this study as well.

All patients with COVID-19 were identified based on WHO interim guidance. The ‘index date’ was the date when the patients were first diagnosed with COVID-19 between January 1^st^,to February 29^th^, 2020. This study was approved by the institutional review boards of all participating medical centers.

### Main Outcome Measures

The main outcomes collected in all hospitalized and confirmed COVID-19 hospital patients were CFR, ICU admission rate, and mechanical ventilation usage after hospital admission. Mortality was defined by death certificates. All COVID-19 patients were followed up to hospital discharge or death at the end of study period (February 29^th^, 2020).The clinical risk factors for ICU admission, mechanical ventilation (both invasive and non-invasive ventilator), and mortality were compared. Patients who did not have these outcomes were categorized as “none.”

### Covariates

All demographic characteristics were determined at index date, including age, gender, race and smoking history. Individual-level data such as exposure history, observation days before hospitalization, the number of symptoms at index date, diagnostic investigations (lab and radiology), all medical procedures, and prescriptions were retrieved from electronic medical records. Comorbidities were also included as clinical covariates.

Clinical symptoms at the index date included fever, nasal congestion, headache, cough, sore throat, sputum production, fatigue, shortness of breath, breathing difficulties, nausea or vomiting, diarrhea, anorexia, abdominal pain, and myalgia or arthralgiachrchan. Lab tests included complete blood count, coagulation profile, serum biochemical tests (renal and liver function, creatine kinase, lactate dehydrogenase, and electrolytes), myocardial enzymes, interleukin-6 (IL-6), and procalcitonin. The comorbidities of patients with COVID-19 included coronary heart disease, diabetes, COPD, hypertension, cerebrovascular disease, chronic kidney disease, chronic liver disease, cancer, and others.

### Statistical Analyses

Continuous and categorical variables are presented as median (IQR) and n (%), respectively. We used the Mann-Whitney U test, χ^2^ test, or Fisher’s exact test to compare differences between survivors and non-survivors where appropriate. Multivariable logistic regression models were conducted to identify factors associated with ICU admission and mechanical ventilation in each cohort, reported as adjusted odds ratios (AOR) and their corresponding 95% confidence intervals (CI).

After patients were diagnosed, the time to death or the time to hospital discharge was examined using the Fine-Gray cumulative incidence functions (CIF). In addition, time to the main outcome – death – was further examined in subgroups of both cohorts by age, gender, comorbidities, and geographic region of residence. Finally, a multivariate Cox proportional hazard model was used to identify factors associated with the cumulative incidence of mortality. Two sensitivity analyses were performed, first by excluding patients with any comorbidity, and second by using multivariate cause-specific hazards models to identify factors associated with the cumulative incidence of mortality in our study. All analyses were conducted using SAS version 9.4 (SAS Institute, Inc., Cary, NC) at a significance level of p<0.05.

## RESULTS

Of the 432 confirmed COVID-19 patients, 9.53% were admitted to the ICU, 27.3% required mechanical ventilation and 33.1% died (Table 1). There were significantly higher proportions of COVID-19 patients of advanced age, more severe disease, and more comorbidities across all three complication outcomes (mortality, ICU admission and mechanical ventilation). In addition, greater proportions of patients who died were current smokers, with symptoms of diarrhea, fatigue, breathing difficulties at the time of diagnosis, and ground-glass or bilateral opacities on chest imaging.

**Table 1.**
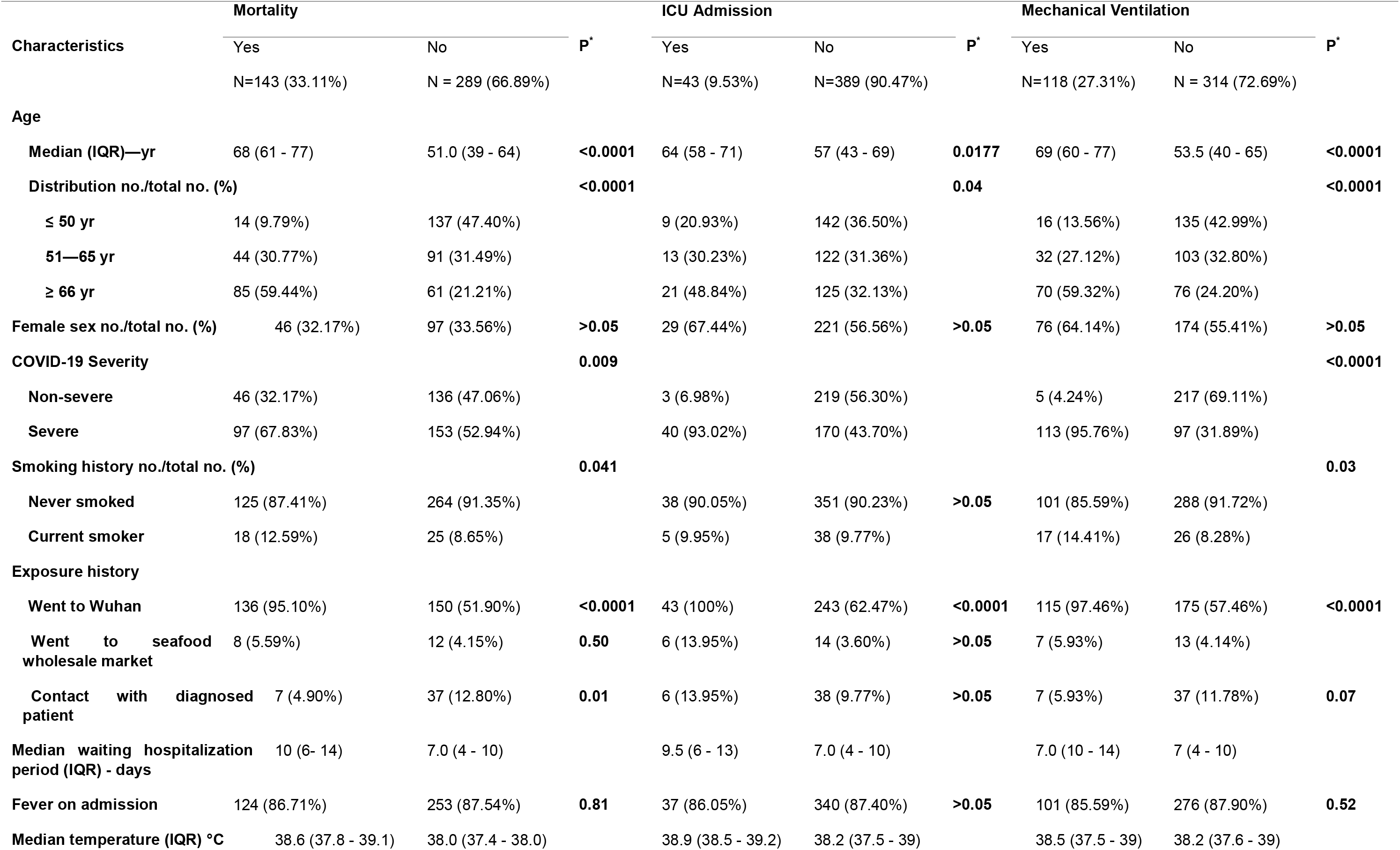
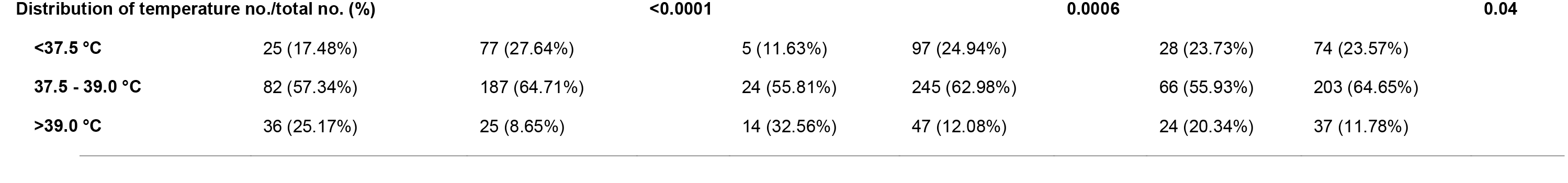
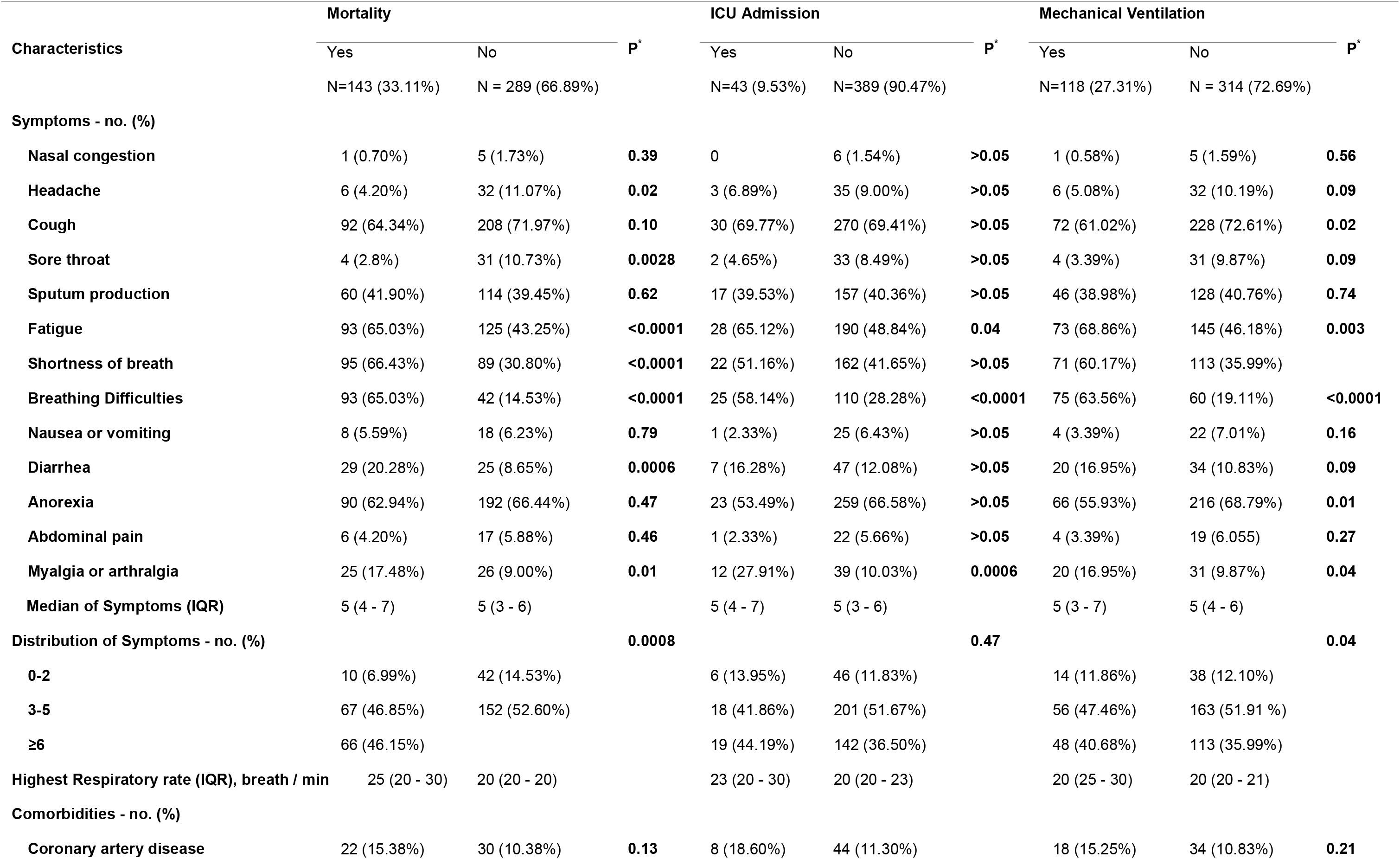
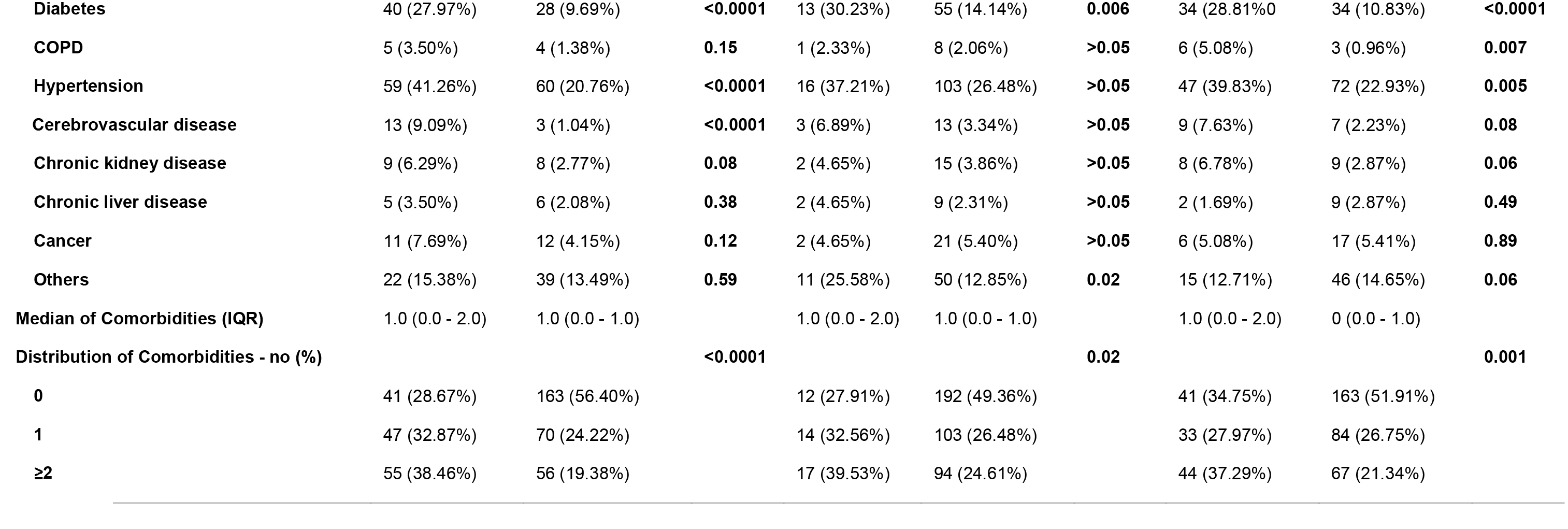
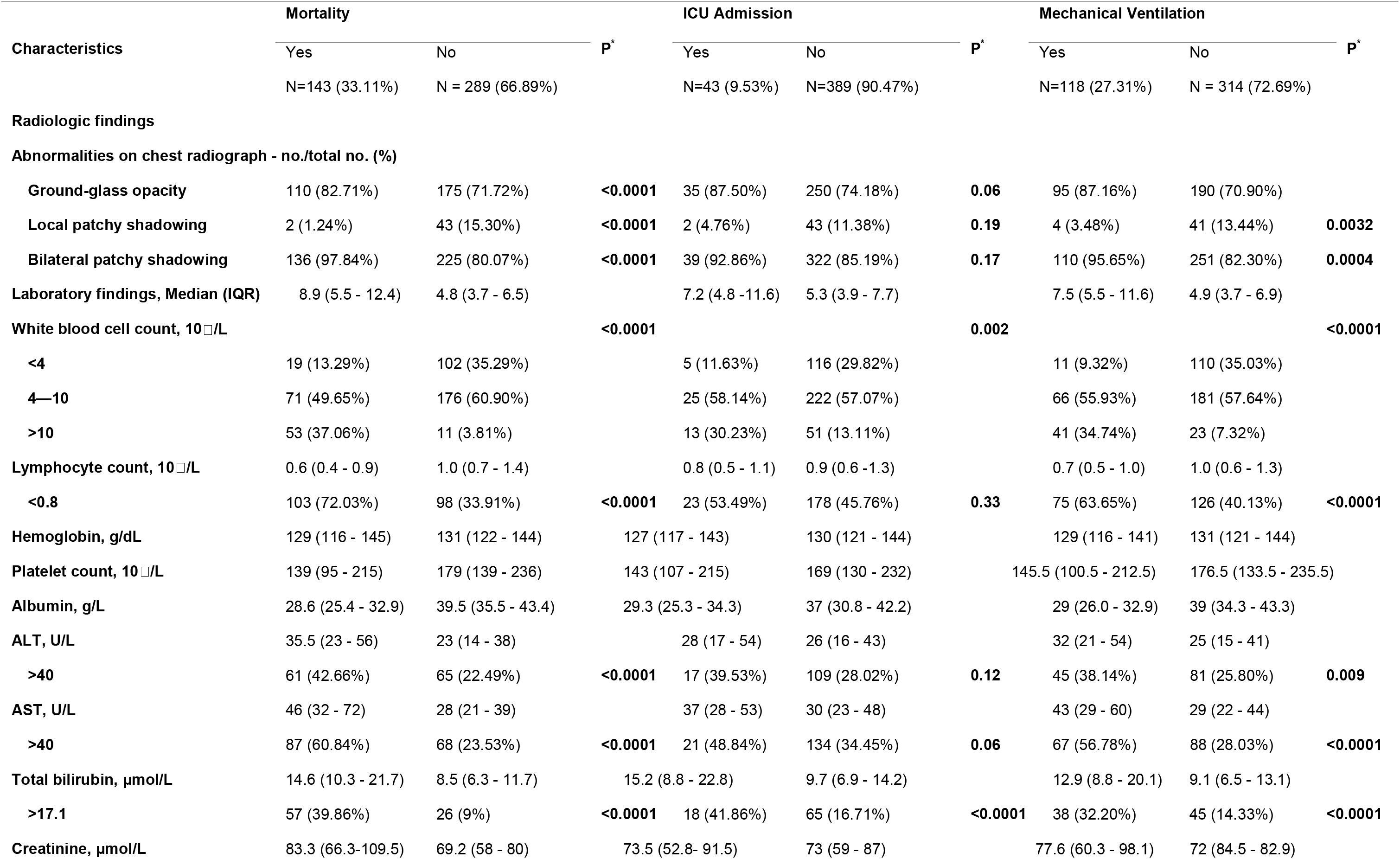
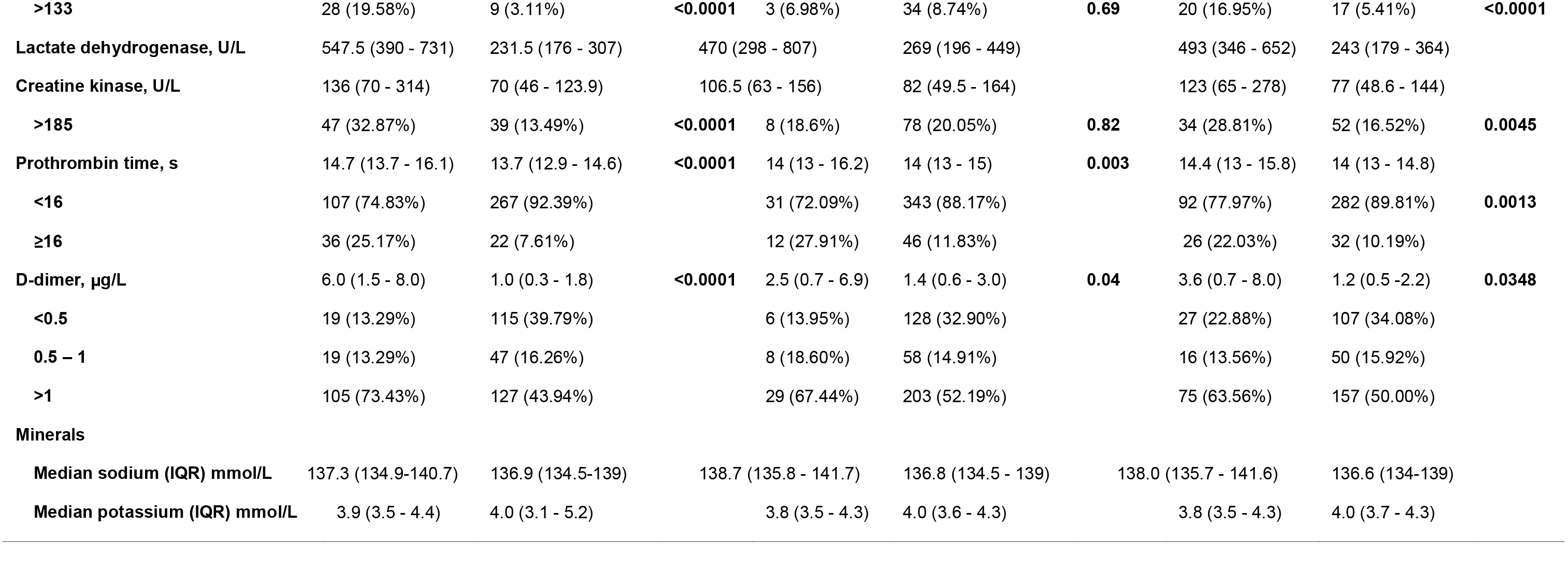
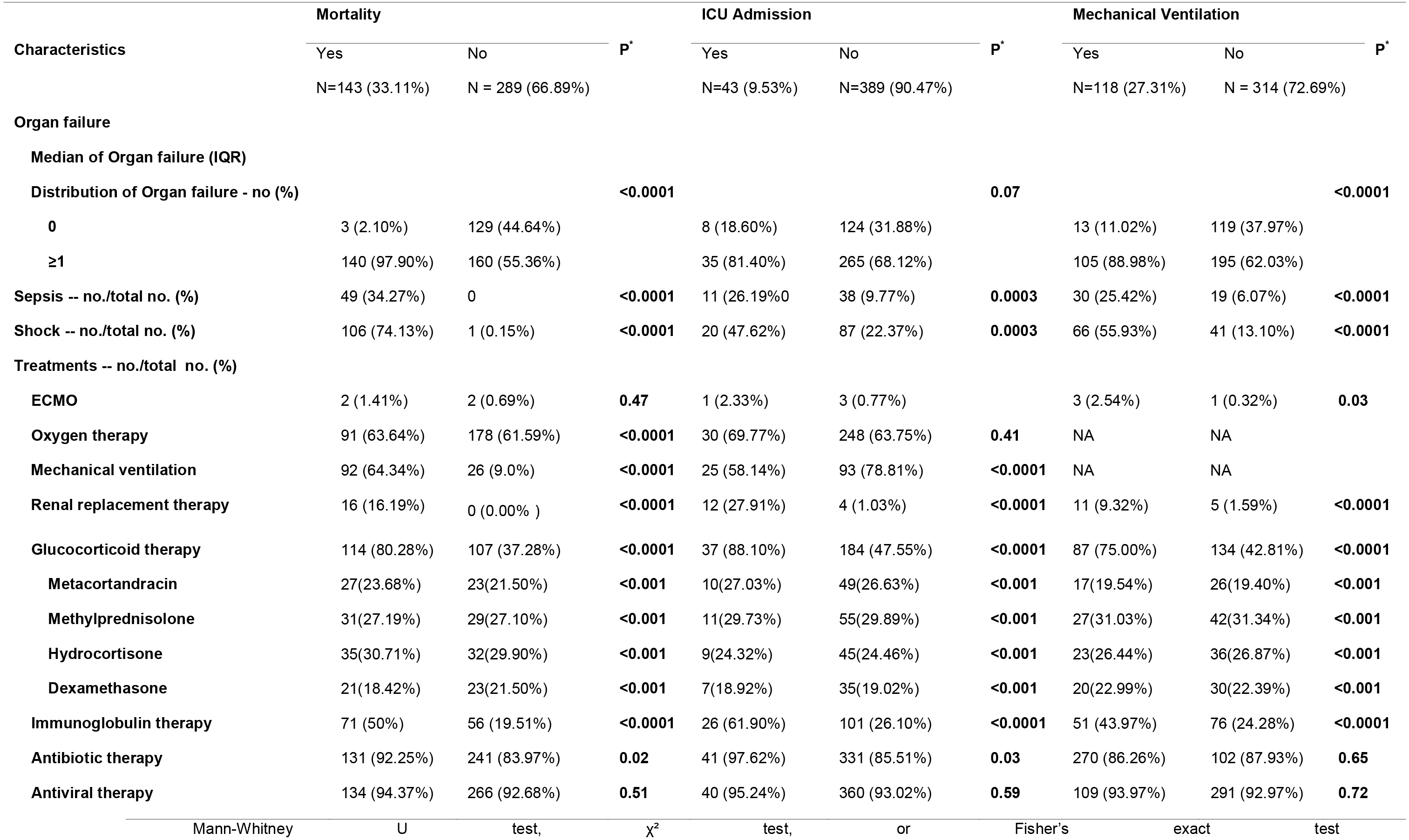
Clinical Characteristics of the All Patients, According to the Mortality, ICU Admission, and Mechanical Ventilation

### Adjusted Cox proportional hazard model for CFR

Table 2 displays the analysis from multivariable Cox proportional hazards models of factors associated with mortality during the 30 days after COVID-19 patients were hospitalized. Age over 65 years (adjusted hazard ratio (HR) 2.09, 95% CI 1.02-4.05), severe disease at index date (HR 5.02, 95% CI 2.05-12.29), current smoker (HR 1.67, 95% CI 1.37-2.02), temperature >39^°^C at index date (HR 2.68, 95% CI 1.88-4.23), multiple comorbidities (diabetes, hypertension, CVA) (HR 2.12, 95% CI 1.62-3.09), bilateral patchy shadowing of chest CT or X-ray (HR 3.74, 95% CI 1.78-9.62), organ failure (HR 6.47, 95% CI 1.97-26.23), high leukocyte count (HR 2.19, 95% CI 1.11-4.31), and a high D-dimer level (HR 1.87, 95%CI 1.87-3.38)] were associated with higher risk of mortality. The following interventions were associated with higher adjusted hazard ratio for mortality: mechanical ventilation (HR 2.35, 95%CI 1.14-4.82), ICU admission (HR 4.92, 95%CI 1.37-17.64), and treatment with glucocorticoids (HR 1.60, 95%CI 1.04-2.30).

**Table 2.**
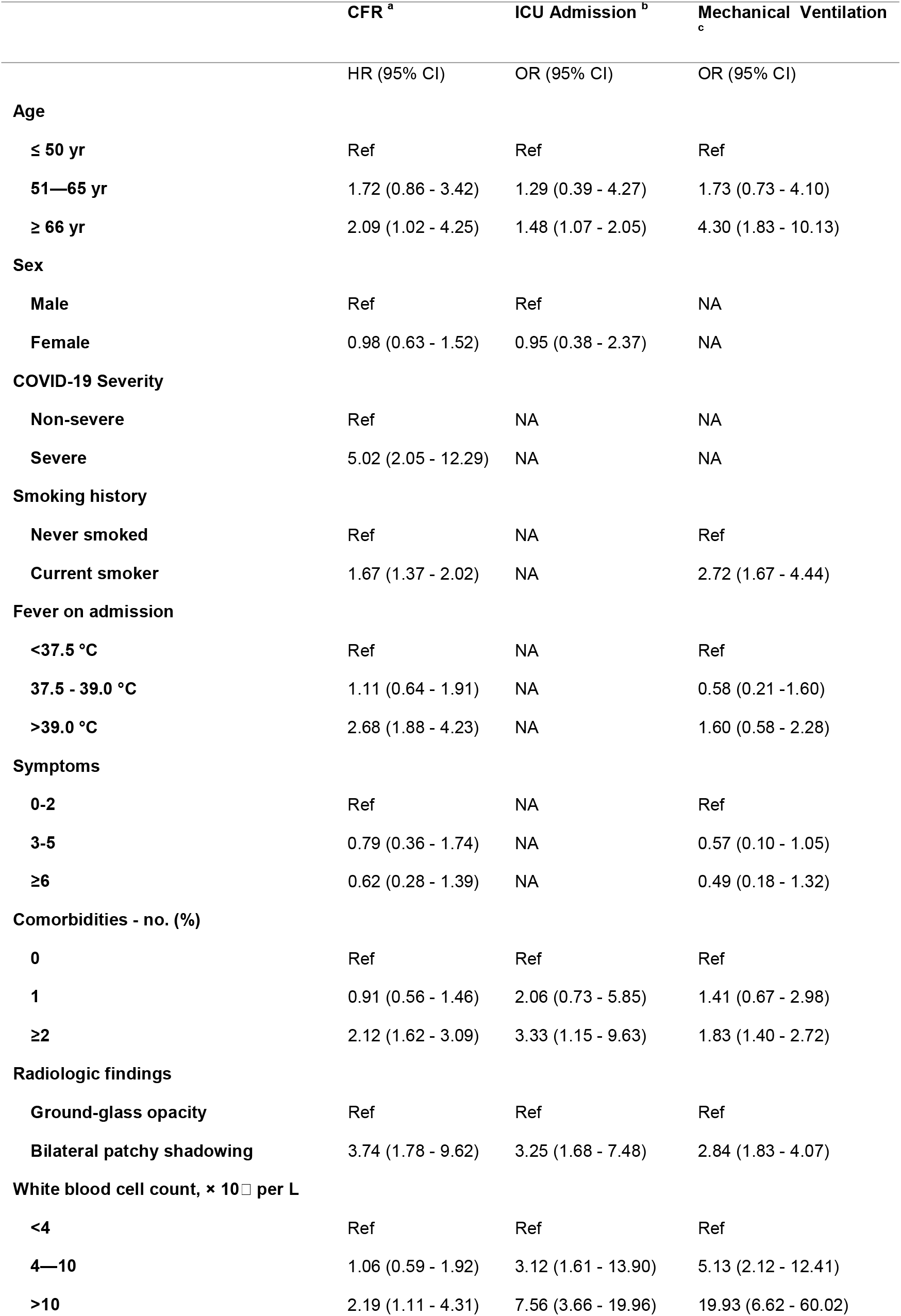
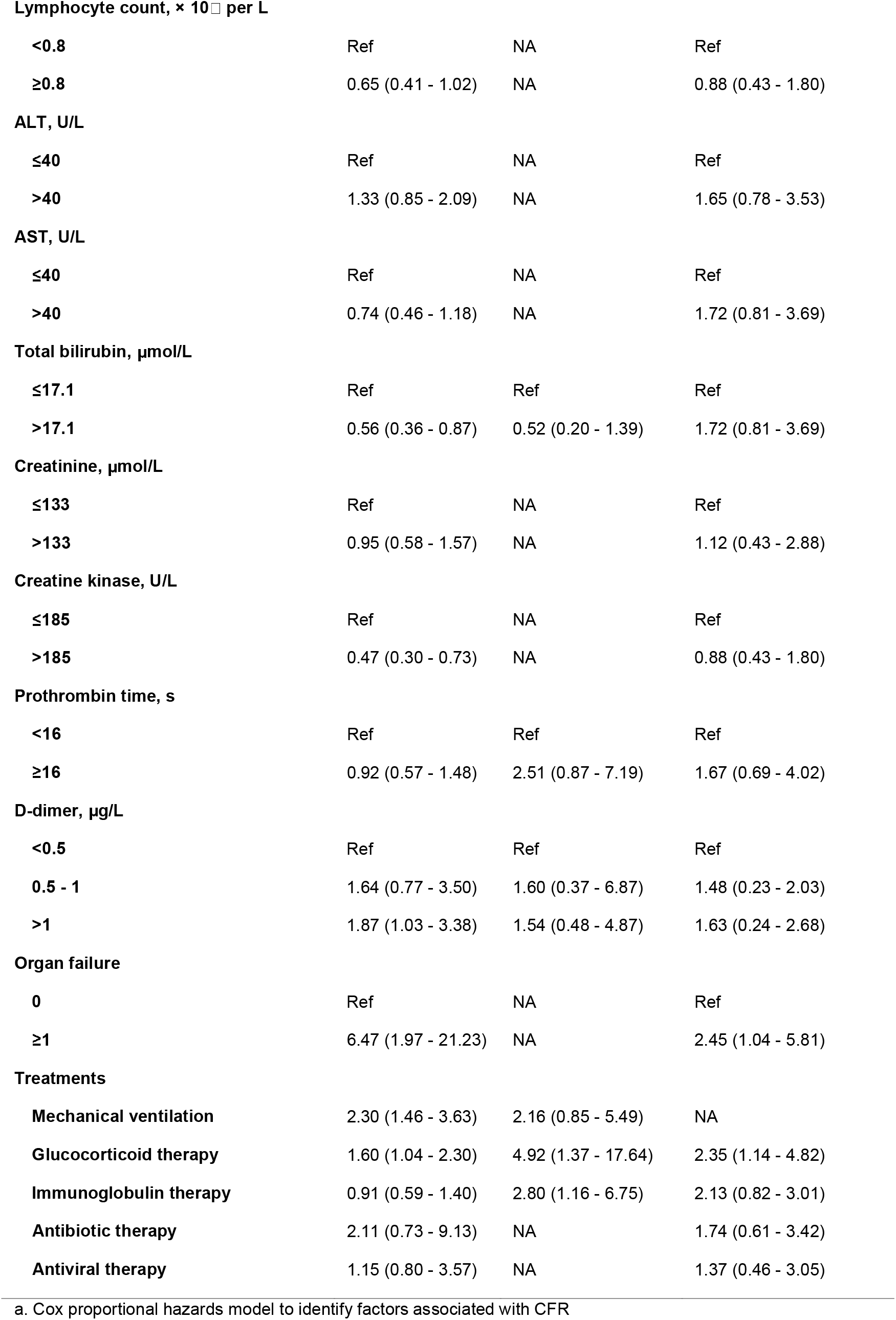
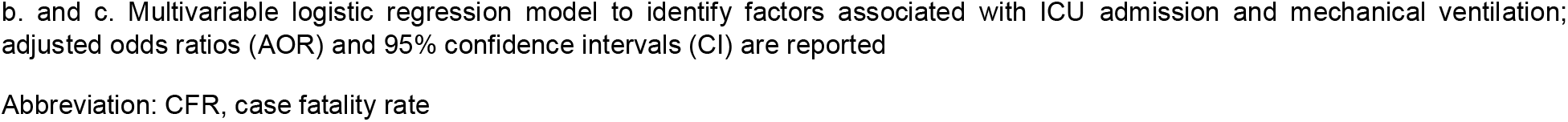
Factors Associated with CFR, ICU Admission and Mechanical Ventilation

### Adjusted Logistic Regression Model for ICU Admission and Mechanical Ventilation

Results from multivariate logistic regression analysis models showed that subjects who were older than 65 years, had more than one comorbidity, or had patchy bilateral shadowing of chest CT or X-ray were more likely to require mechanical ventilation or be admitted into the ICU (Table 2). Similarly, patients who were older than 65 years, current smokers, had more than one comorbidity, organ failure, or a leukocyte count ≥4 × 10^9^ per liter were more likely to require mechanical ventilation (Table 1).

Sensitivity analyses using cause-specific hazard models for the 432 hospitalized patients found similar results for mortality. After excluding those patients with comorbidities, the risk factors associated with mortality and mechanical ventilator support were consistent with the primary findings. Due to the small number of patients admitted to ICU, sensitivity analysis was not possible in this subgroup.

## DISCUSSION

This multi-center retrospective cohort study of 432 patients with confirmed COVID-19 admitted to three academic medical centers in Hubei province, China, from Jan 1st – Feb 29th, 2020, shows that 33.1% patients died within 30 days after COVID-19 diagnosis. The risk of death in hospitalized patients is far greater than that of the total population diagnosed with COVID-19, since overall CFR include both hospitalized and non-hospitalized cases, and the majority of confirmed cases of COVID are less severe and do not require hospitalization. As of May 5, 2020, there were 57,682 COVID-19 cases and 3130 COVID-19 related deaths in Hubei province. The reported CFR across Hubei province is 5.1%, compared to a global CFR of 4.1%.

The difference in CFR may be in part due to the fact that Union Hospital medical center admitted more elderly patients than others during the the earlier period of the COVID-19 outbreak in China. This is supported by our findings in this study that positive COVID-19 patients who were older than 65 years old admitted to hospitals is associated with a 2-fold increased risk of death after 30 days.

In contrast to a study by Zhou et al of inpatients at different hospitals in Wuhan,^11^ our study identifies a number of other factors associated with increased risk of death, ICU admission, and mechanical ventilation that may be helpful for clinicians assessing prognosis and risk cohorting. The higher risk of death and mechanical ventilation in current smokers is a notable finding, and may be relevant in countries where smoking is common. It may represent a potentially modifiable risk factor, and a may serve as an impetus to encourage smoking cessation or develop smoking prevention programs to mitigate risks during coronavirus outbreaks. In addition, our study suggests that patients who present to hospital with increased age, multiple comorbidities, higher temperature, higher leukocyte count, higher D-dimer, organ failure, and radiologic abnormalities should be assessed as a cohort at higher risk of requiring ICU, ventilatory support and death.

It is also notable that diarrhea was proportionately higher in patients who died during hospitalization (Table 1), however, this symptom was not individually assessed in the adjusted proportional analysis, and remains to be explored in future analyses. A recent study of 204 patients with confirmed COVID-19 in Hubei found that 48.5% of patients presented to hospital with gastrointestinal symptoms as their chief complaint, although the association with death was not explored.^15^

The association of higher D-dimer levels (> 1 μg/mL) and bilateral opacities on chest imaging with adverse outcomes and death is in agreement with previous studies of Severe Acute Respiratory Syndrome CoV (SARS-CoV) and Middle East Respiratory Syndrome CoV (MERS-CoV) that described massive inflammatory cell infiltration within the lower airway^16^ leading to pro-inflammatory cytokine/chemokine storm.^17^ This rapidly evolving immunological dysfunction or dysregulation can lead to acute lung injury (ALI) and ARDS. The 2019 novel coronavirus (SARS-CoV-2) has a similar genome, and may share similar pathophysiologic findings, although SARS-CoV may cause even more severe cytokine storm resulting in its higher CFR. Increased D-dimer concentrations among patients with pneumonia have been observed in studies due to increased coagulation activity.^18^ Microvascular thrombosis can result from excessive netosis observed in COVID-19 patients.

Although activation of the inflammatory cascade appears to contribute to adverse outcomes, we found that corticosteroids were associated with a higher risk of death, ICU admission, and mechanical ventilation. In 2003 and 2004, corticosteroids were widely used to treat SARS, first in mainland China and then in Hong Kong and beyond. The hypothesis of using corticosteroids was to reduce “cytokine storm” related acute lung injury, resulting from release of early response cytokines such as interferon-gamma (IFN-γ), tumor necrosis factor (TNF-α), interleukin 1 (IL-1), and interleukin 6 (IL-6).^17,20^ However, a number of observational studies have shown an association of corticosteroids with poorer outcomes in COVID-19 and the balance of evidence questions their use for treatment of human coronaviruses including SARS-CoV, MERS-CoV, and COVID-19.20^19,20^ The reasons for the adverse association could be that (1) corticosteroids are harmful overall given the net clinical impact on other unrecognised physiologic mechanisms, despite reducing the inflammatory response, or (2) more severely affected patients are more likely to be prescribed corticosteroids and die, and selection biases remain in our mortality estimates even after calculating adjusted hazards ratios. These results emphasize the importance of the data from the RECOVERY (Randomised Evaluation of COVid-19 thERapY) trial (www.recoverytrial.net/), which found that dexamethasone reduced the risk of 28-day mortality. Our study is strictly not a direct comparison to that of RECOVERY, and our results actually highlight just how important it is to have a controlled trial surrounding the ‘when’ and the ‘to whom’ one should commence / administer corticosteroid treatment. Current recommendations are to use low-dose corticosteroid therapy in patients with COVID-19 and refractory shock^13^.

In line with previous studies of SARS-CoV and MERS-CoV,^21,22^ our findings also provide evidence for higher risk of death with increasing age in hospitalized patients with COVID-19. In addition to the higher risk of mortality in elderly patients, we also found they were more likely to be admitted into ICU and more likely to require mechanical ventilation. We also found a higher likelihood of mortality in patients with multiple morbidities, such as coronary heart disease, which has previously been associated with poor outcomes of respiratory viral infections, and diabetes, which has previously been linked to reduced immune system function.^23-25^

Other interesting findings were that a higher proportion of patients with positive exposure history presented with more severe disease. The reason for this association is unclear.

### Strengths and Limitations

Of the currently published COVID-19 observational studies on mortality to date, the sample size of our study is among the largest. Despite that, it may still lack sufficient power to determine true associations in our analysis. Furthermore, the treatment regimens across different centers may differ, including ICU admission criteria, mechanical ventilation criteria and modalities, use of antibiotics, antivirals, and other supportive interventions. Considering the retrospective observational nature of our study, it is difficult to eliminate selection biases and remaining confounders. For this reason, conclusions regarding the risk-benefit tradeoffs for interventions including corticosteroids, antibiotics, antivirals, mechanical ventilation, and ICU admission cannot be made from this analysis. However, this analysis is the best we can provide to enlighten current outcomes related to approaches to treatment for COVID-19 that were utilized in Hubei hospitals in early 2020. Due to subgroup limitations for each comorbidity, we combined all comorbidities together for analysis. Larger studies are needed to further elucidate which patients are at most risk of death, ICU, or require mechanical ventilation by specific co-morbidities. Finally, this was a retrospective case series study that relied on abstracting data from clinical charts. Accordingly, information was limited to that provided in the charts at the time of patient care.

## CONCLUSION

In hospitalized patients with confirmed COVID-19 in Hubei, the need for mechanical ventilation and risk of death was high. A number of patient-level demographics were associated with increased risk of mortality, including age over 65 years, current smoker, diabetes, hypertension, and cerebrovascular disease. Clinical characteristics associated with mortality included temperature >39^°^C, diarrhea, fatigue, shortness of breath, breathing difficulties, higher disease severity, organ failure, and ground-glass opacity or bilateral patchy shadowing on chest imaging. Risk of mortality was also associated with mechanical ventilation and ICU admission. Corticosteroid treatment was associated with increased mortality. These results underscore the urgency of adequate prospective clinical trials to inform the role of, in particular, corticosteroids, but also mechanical ventilation and ICU admission, in managing COVID-19. Results from the RECOVERY trial have strongly indicated that timing of corticosteroid intervention is key. Knowledge of baseline demographic risk factors and clinical predictors for adverse outcomes may assist clinicians in optimally triaging patients presenting to hospital with COVID-19 during this pandemic. Finally, our study’s results are in line with those results coming from studies published in the West^26-28^.

## Data Availability

Availability of data and material: The corresponding author had full access to all the data in the study and had final responsibility for the decision to submit for publication.

## Declarations

## Availability of data and material

The corresponding author had full access to all the data in the study and had final responsibility for the decision to submit for publication.

## Code availability

Not applicable.

## Authors’ contributions

All authors made a significant contribution to merit authorship, either by writing and editing the manuscript text, and/or analyzing data, and each has seen and approved the final version. HR, FZ, JM, PY, QH, collected the epidemiological and clinical data. HR, JM, DC summarized all data. HR, DW drafted the manuscript. HR, LBS, KB, XG, DW, NH, JS, DC edited the final manuscript.

